# The use of facemasks by the general population to prevent transmission of Covid 19 infection: A systematic review

**DOI:** 10.1101/2020.05.01.20087064

**Authors:** Madhu Gupta, Khushi Gupta, Sarika Gupta

## Abstract

**Background:** The pandemic of COVID-19, caused by severe acute respiratory syndrome coronavirus 2 (SARS-CoV-2), has become a serious worldwide public health emergency. This systematic review aims to summarize the available evidence regarding the role of face mask in community settings in slowing the spread of respiratory viruses such as SARS-CoV-2.

**Methods:** The preferred reporting items for systematic reviews and meta-analyses (PRISMA) guidelines were used for this review. Literature search using PUBMED, Google Scholar and Cochrane database was performed using Medical subject heading (MeSH) words from the year 2000-2020. The articles focused on the use of masks and N95 respirators in healthcare workers were excluded.

**Results:** A total of 305 records were identified, out of which 14 articles were included in the review based upon quality and eligibility criteria. All the articles mentioned about role of face masks in preventing the spread of respiratory viruses like influenza, SARS and SARS-CoV-2, in the community or experimental setting. Studies also suggested that early initiation of face mask usage was more effective. Masks were also reported to be more effective in viruses which transmit easily from asymptomatic individuals, as is now known in SARS-CoV-2.

**Conclusion:** Theoretical, experimental and clinical evidence suggested that usage of face masks in general population offered significant benefit in preventing the spread of respiratory viruses especially in the pandemic situation, but it’s utility is limited by inconsistent adherence to mask usage.

## Introduction

The ongoing pandemic of coronavirus (COVID-19), caused by severe acute respiratory syndrome coronavirus 2 (SARS-CoV-2) has become a serious worldwide public health emergency. The number of infected patients is increasing exponentially with every passing day with over two million cases reported worldwide. In the absence of clearly defined therapeutic agents and vaccines, the mitigation of the pandemic and “flattening the curve” is primarily being attempted by nonpharmaceutical interventions, like social distancing and hand hygiene. The usage of face masks in the general population has been a controversial subject as the WHO and most European guidelines did not support the use of facemasks by healthy people in the community. WHO guidelines recommend the use of facemasks only for healthcare workers, symptomatic people and their caregivers^1^. On the other hand, few countries like Japan and Hong Kong have recommended the use of masks by the general population ^2,3^. These countries have also been relatively more successful in reducing the spread of the infection, better than Europe and USA. In view of these conflicting guidelines, we conducted a systematic literature review to assess the utility of facemasks in the general population for reducing the spread of respiratory viruses like COVID 19, especially in the setting of a pandemic.

## Material and methods

A systematic review of published literature from the year 2000-2020 was done, to estimate the effectiveness of face masks (surgical/cloth) in community settings, in reducing the spread of respiratory viruses. The preferred reporting items for systematic review and meta analysis (PRISMA) guidelines were used for the review. The publications chosen for this study included randomized control trials, non randomized experimental studies and observational studies. The articles excluded from the study were the ones that focused on the use of masks and N95 respirators in healthcare workers.

## Literature search in data sources

Pubmed and Cochrane databases were used for searching the literature, using the following MeSH words - *‘face masks’ or ‘ masks’ or ‘surgical masks’ or ‘cloth masks’ and ‘respiratory infection’ or ‘respiratory virus’ or ‘influenza’ or ‘SARS’ or ‘Subacute respiratory syndrome’ and ‘COVID 19’ or ‘coronavirus’, ‘influenza pandemic’ and ‘community’ or ‘household’*. We used an open date strategy up to April 2020. Considering that studies published in local journals might not be indexed on fore-mentioned databases, we made an additional search on Google Scholar using the same keywords. We set a limit of 30 results per combination of search words ranked on the basis of ‘Relevance’.

### Screening

The article titles, abstracts and summary of all the studies were screened for potential inclusion in this review. The authors then reviewed the full text versions of the selected articles to determine inclusion. Those that did not focus on use of masks for respiratory infections by healthy individuals in household or community settings were excluded to arrive at the final list. The full texts of these articles were then studied independently by three investigators.

### Data extraction and analysis

The data from the articles was extracted by the authors, the key points were assessed and noted from each selected article. Because the designs of these studies, nature of participants, interventions and reported outcome measures varied significantly, we focused on describing the studies, their results and their limitations and on qualitative assessment rather than meta-analysis. The findings from the key experimental articles were then tabulated and summarized.

## Results

A total of 305 records were identified through appropriate databases; 53 were removed as they were duplicates and 145 were removed after the readers screened through the title and abstract. Full texts of the remaining 107 articles were read and 76 were removed on the basis of pre-defined exclusion criteria. Seventeen of the remaining 31 articles were excluded due to quality issues leaving a final 14 studies in this review. (Figure 1).

**Figure 1.**
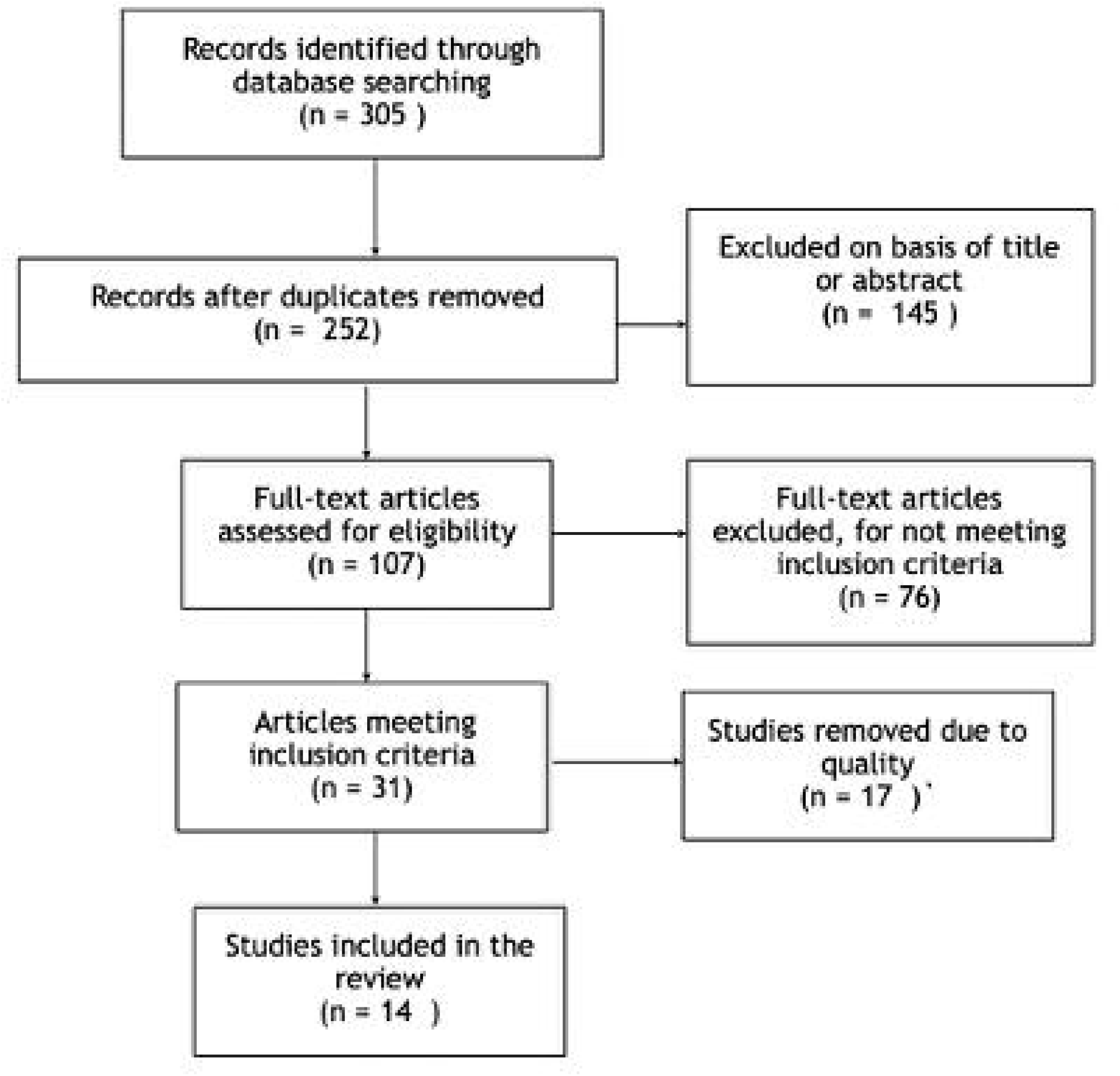
Flow Diagram for Search Strategy:

The summary of the studies reviewed in this review is given in table 1, and described below:

**Table 1.**
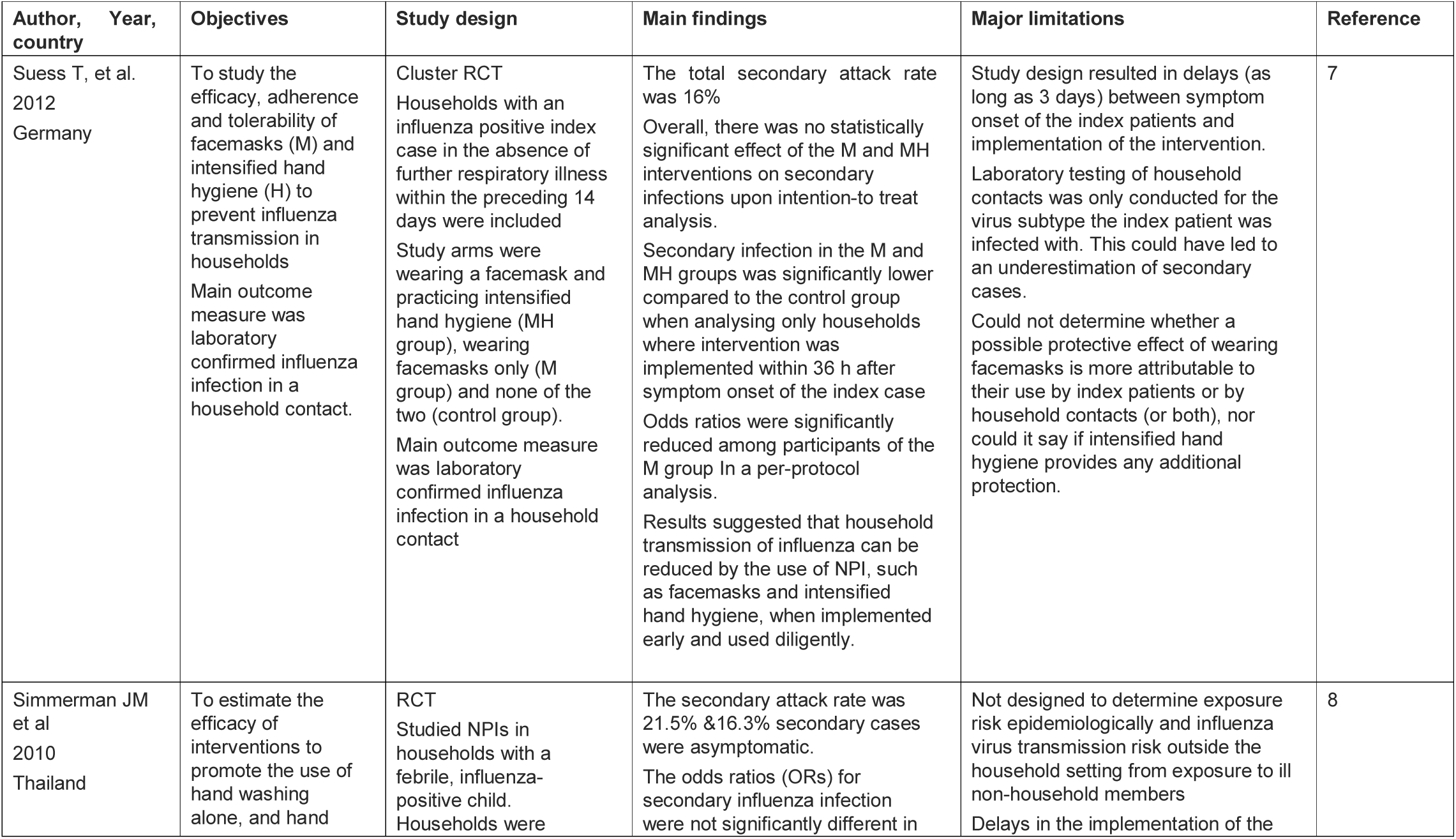
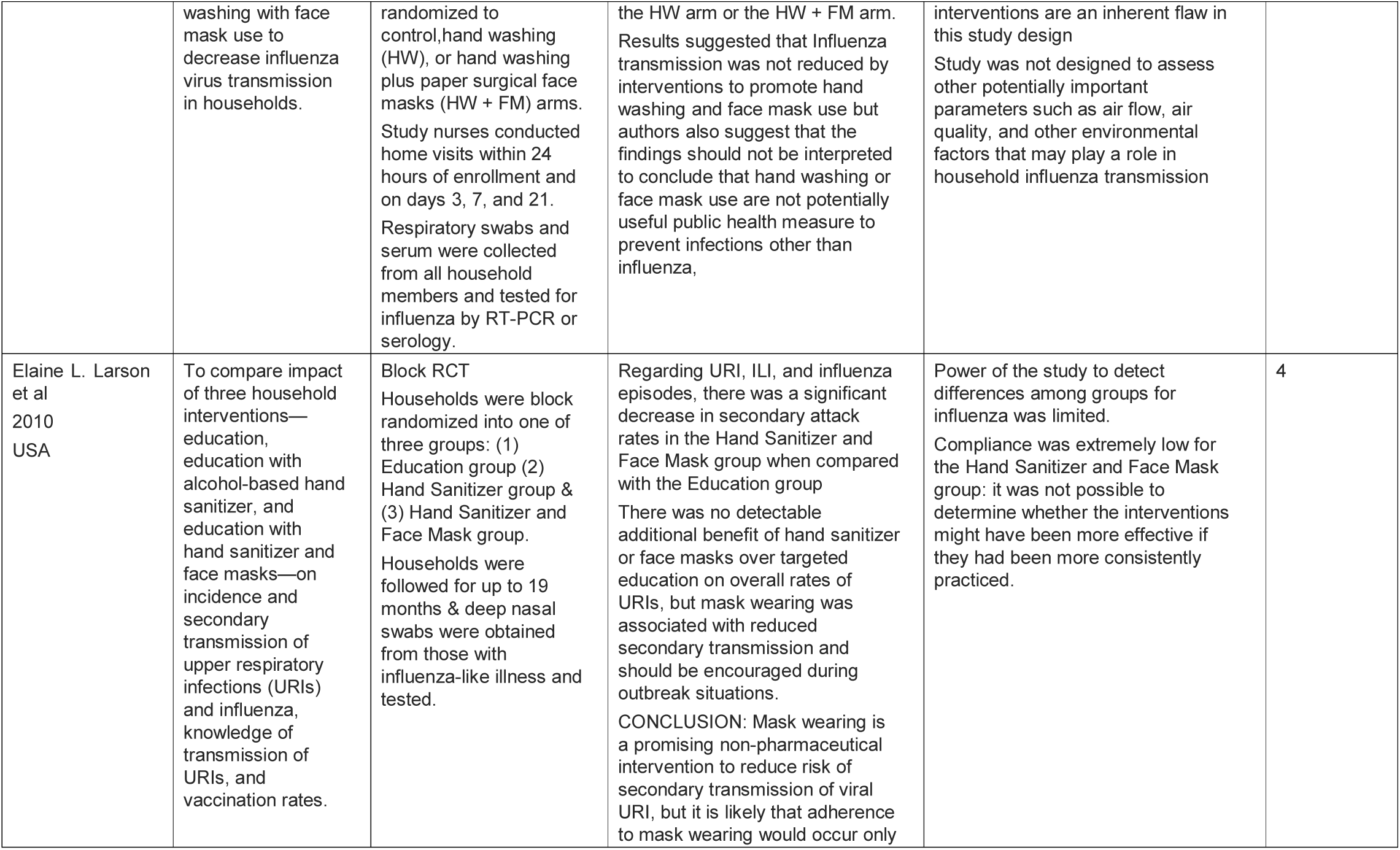
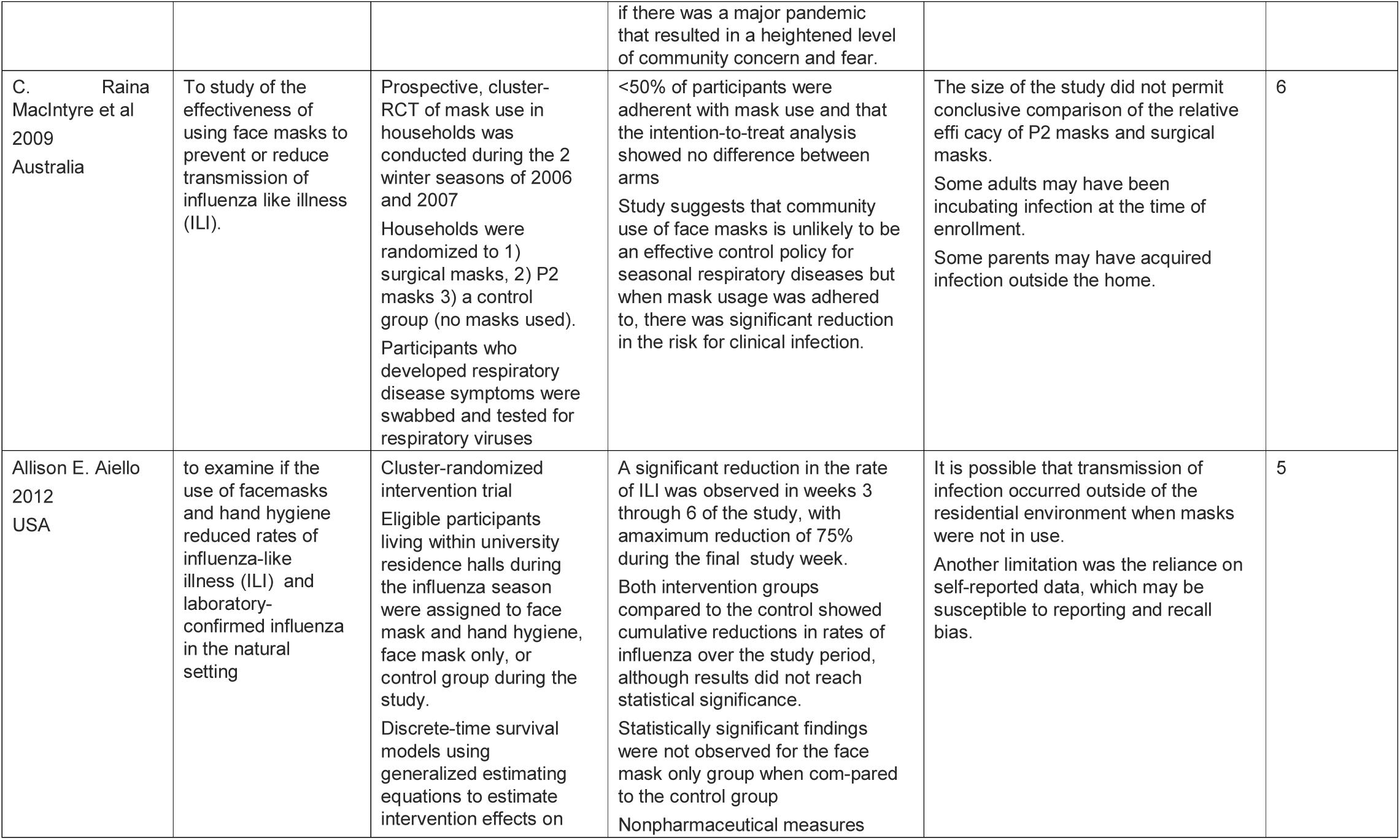
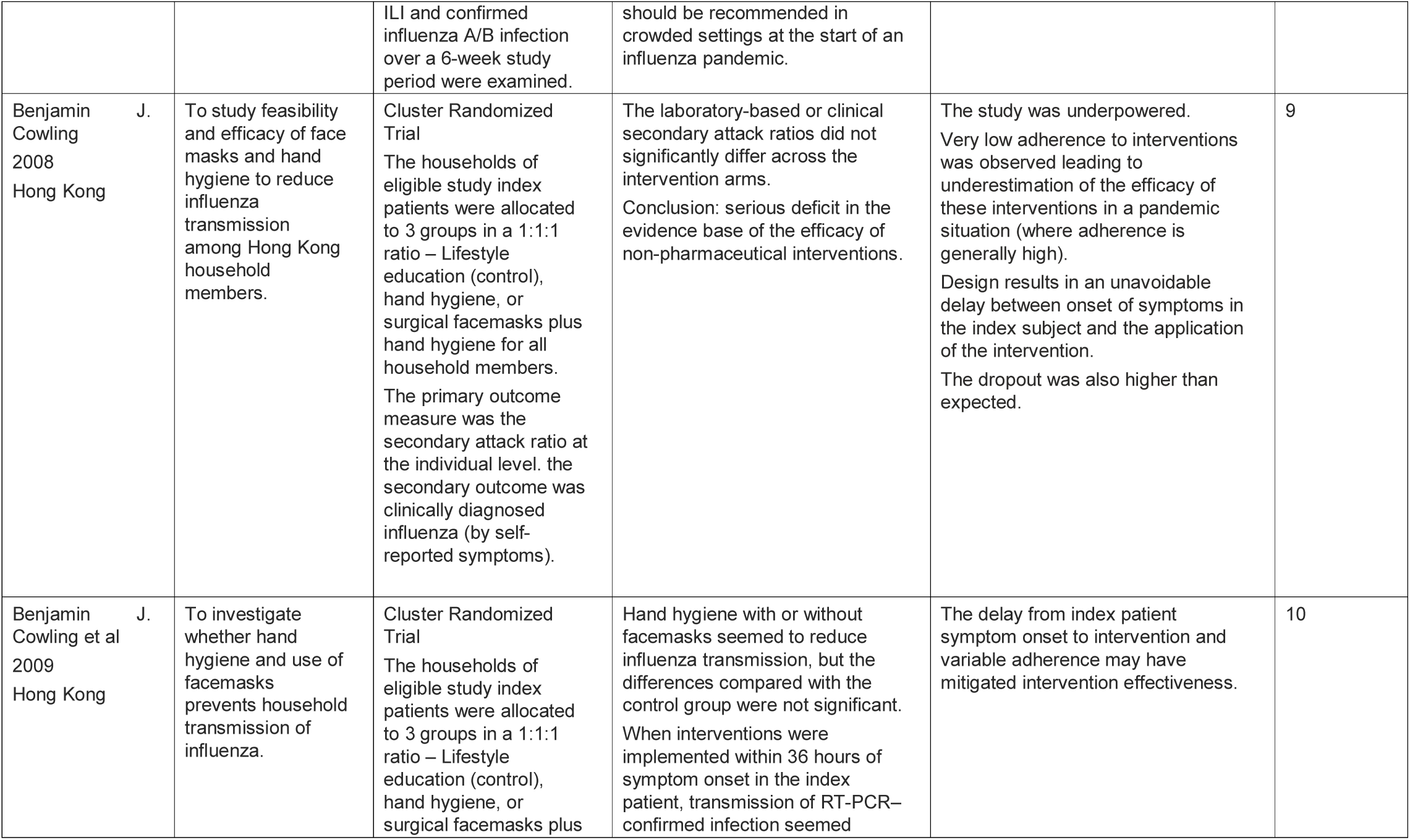
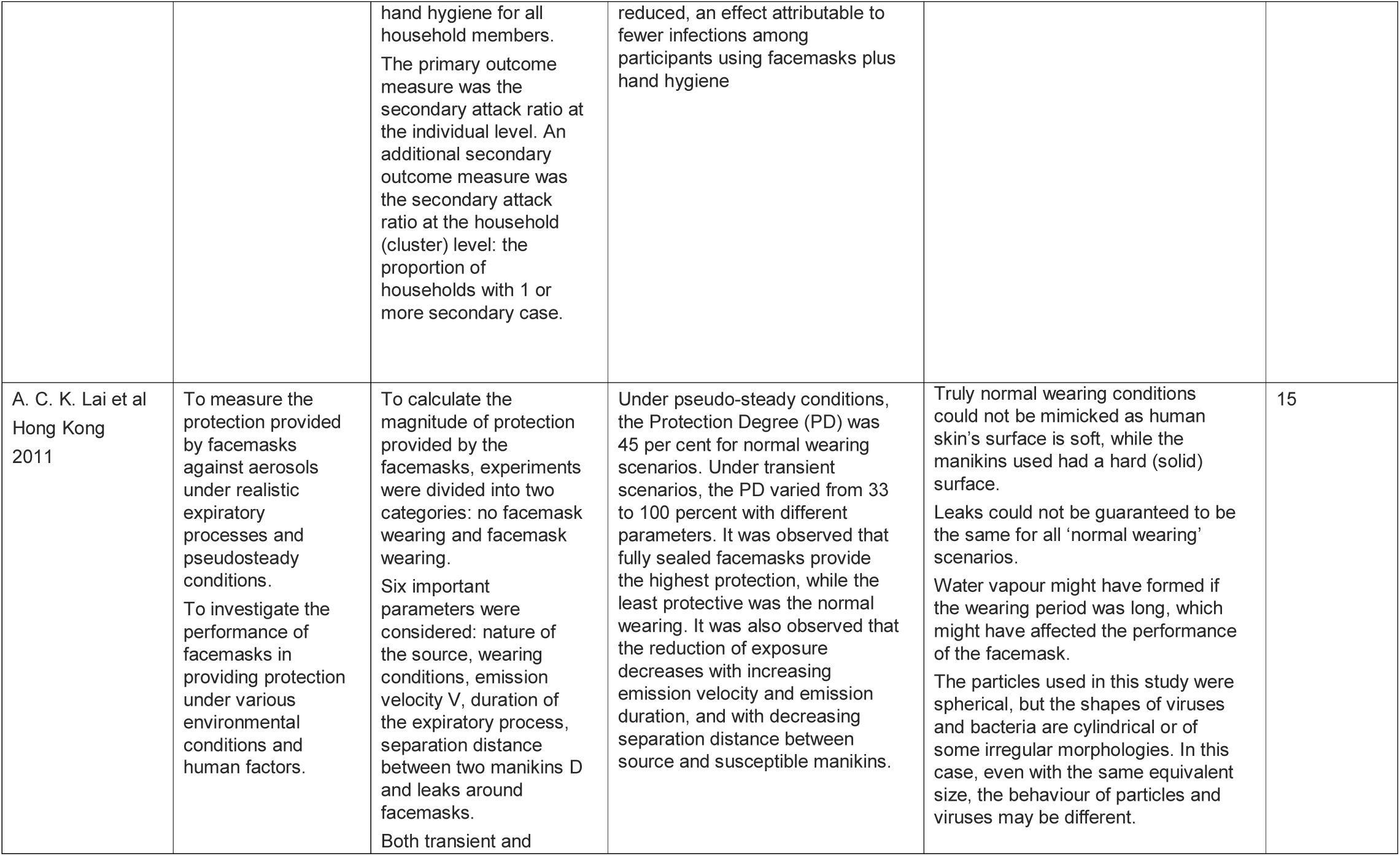
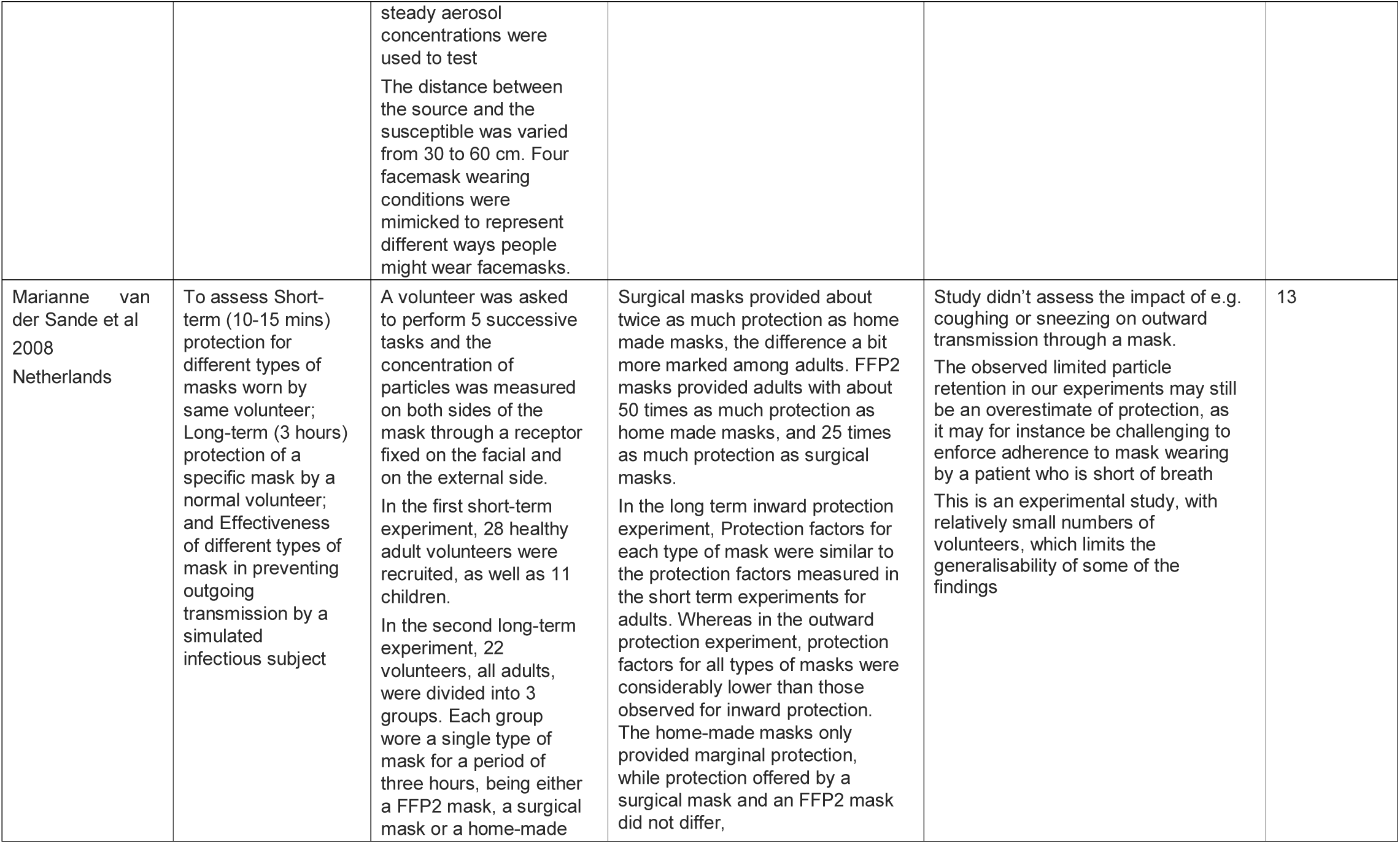
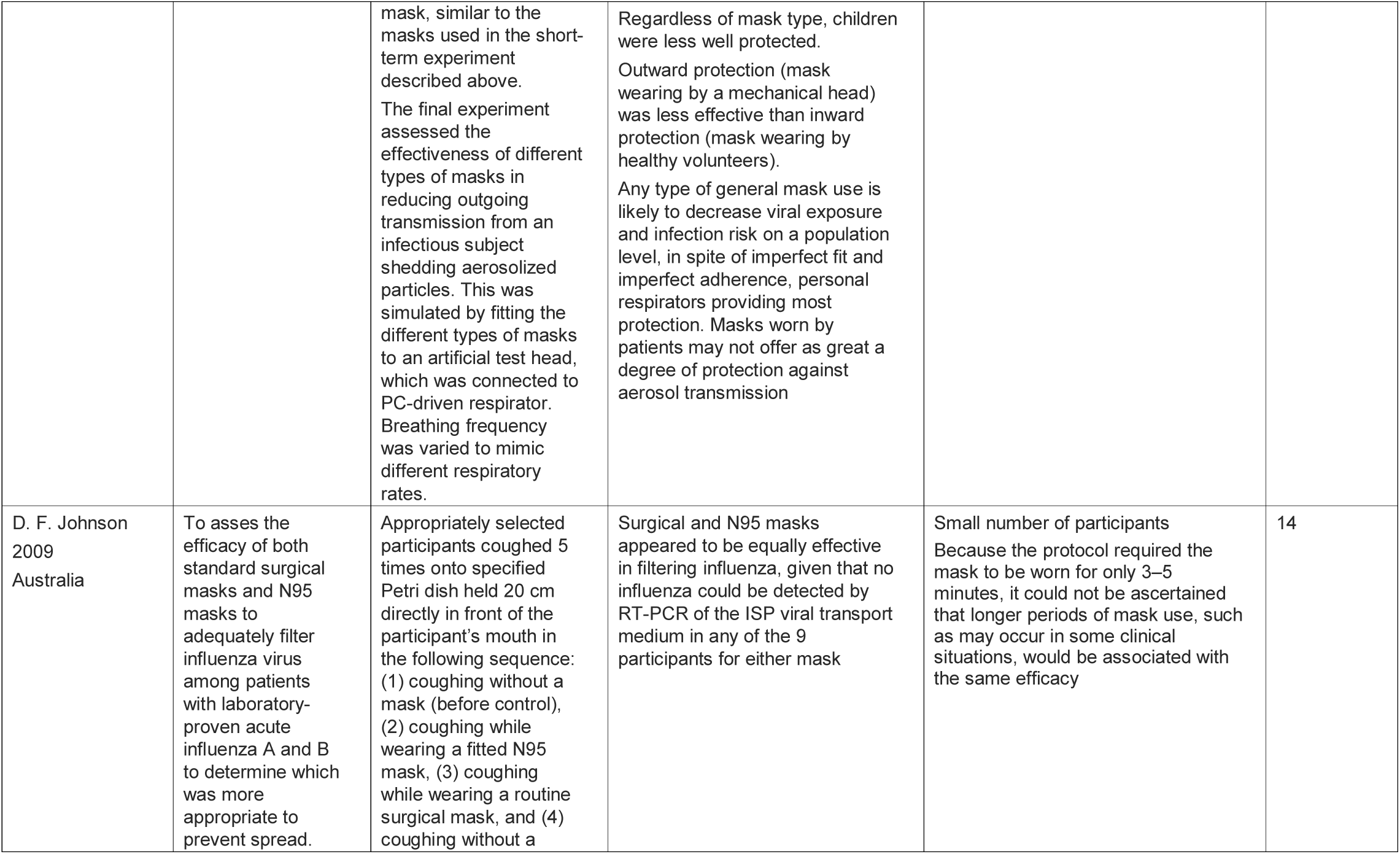
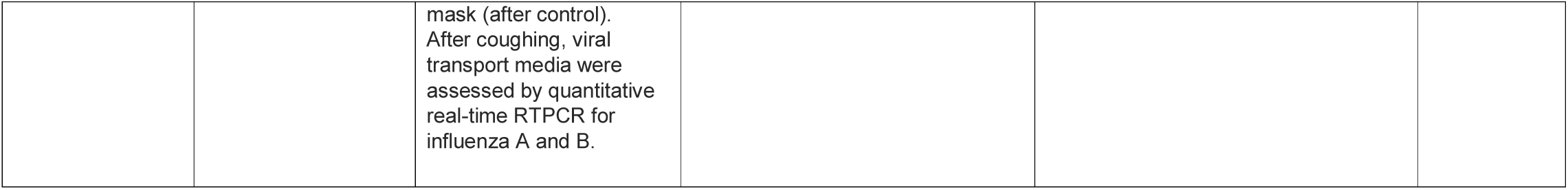
Summary of studies reviewed for use of face masks.

## Experimental studies

### Randomized controlled trials

Seven randomized controlled or cluster randomized trials were done in community settings. A trial conducted amongst 509 primarily Hispanic households found that mask wearing was associated with reduced secondary transmission of respiratory viruses / influenza-like illness/ laboratory-confirmed influenza and should be encouraged during outbreak situations. Use of hand sanitizer alone resulted in no significant reduction in the outcome. The masks used were regular surgical masks and not National Institute for Occupational Safety and Health certified N95 respirators.^4^ Another randomized intervention trial was conducted amongst 1437 young adults, living in university residence halls during the 2006–2007 influenza season. The authors found a significant reduction in the rate of Influenza like Illness (ILI) among participants randomized to the face mask and hand hygiene intervention during the latter half of this study, ranging from 35% to 51% when compared with a control group that did not use face masks.^5^ There was another prospective, cluster-randomized trial of mask use in households, conducted during the 2 winter seasons of 2006 and 2007 in Sydney, Australia. The key findings of this study were that <50% of participants were adherent with mask use. However, adherent mask users (P2 or surgical) in the study showed a relative reduction in the daily risk of acquiring a respiratory infection in the range of 60%–80%^6^. A randomized control trial done in Berlin, Germany in 2009-2011 showed encouraging results with early implementation of face mask use and hand hygiene in preventing spread of influenza in households^7^. Three other randomized mask intervention studies done in Bangkok and Hong Kong, reported no significant reduction in secondary transmission of ILI.^8,9,10^

### Non Randomized controlled trials

Van der saande et al, (2008), conducted experiments with healthy volunteers to assess short-term and long-term protection provided by different types of masks worn during 10-15 minutes by the same volunteer following a standardized protocol. Inward protection was defined as the effect of mask wearing to protect the wearer from the environment; outward protection was defined as the effect of a mask on protecting the environment from the generation of airborne particles by a patient (or in this case a mechanical head).^13^ The study result revealed that all types of masks provided protection against transmission by reducing exposure during all types of activities. The inward protection of a cloth mask was approximately 2.5 times more than no mask. Surgical masks provided about twice as much protection as homemade masks. In the final experiment, they also assessed the effectiveness of different types of masks in providing outward protection or reducing outgoing transmission from an infectious subject shedding aerosolized particle. This was simulated by fitting the different types of masks to an artificial test head, which was connected to PC-driven respirator. The home-made masks only provided marginal protection, compared with no mask. The outward protection offered by a surgical mask and a FFP2 (European equivalent of N95 mask) was only 2-3 times, compared to a no mask state.^13^

Johnson et al, (2009), did an experimental study to test the performance of surgical and N95 masks to filter influenza virus in nine volunteers with confirmed influenza A or B virus infection. Participants coughed five times onto a Petri dish containing viral transport medium held 20 cm in front of their mouth.^14^ The experiment was repeated with subjects wearing a surgical mask, and wearing an N95 respirator. While influenza virus could be detected by RT–PCR in all nine volunteers without a mask, no influenza virus could be detected on the Petri dish specimens when participants wore either type of face mask. A limitation was that the study did not consider the role of leakage around the sides of the mask.^14^ A study done by Lai et al, (2012), showed that wearing a mask reduces exposure to air borne disease by an average of 45%.^15^ The degree of protection varies between 33-100% depending on the fit of the mask and variables of transmissibility of the infectious agent.^15^

## Observational studies

A matched case-control study was done during the Beijing outbreak of SARS in 2003, among a sample of patients who had no reported contact with other SARS patients. This observational study concluded that people who always wore masks had a 70% lower risk of being diagnosed with clinical SARS compared with those who never wore masks, and persons with intermittent mask use had a 60% lower risk.^11^

Another observational study suggested that, during the height of the SARS epidemic of April and May 2003 in Hong Kong, adherence to infection control measures was high with 76% of the population wearing a face mask The authors found that distributing masks during seasonal winter influenza outbreaks is an ineffective control measure characterized by low adherence, results indicate the potential efficacy of masks in contexts where a larger adherence may be expected, such as during a severe influenza pandemic or other emerging infection.^12^

## Mathematical models

Mathematical models were not collected in a tabular form. Mathematical models have been used to analyze the effectiveness of facemasks in reducing the spread of novel influenza A (H1N1) virus.^16^ The model in this study is a Susceptible-Exposed-Infectious-Recovered (SEIR) model. The conclusion of this study was that the effectiveness of surgical masks is low and therefore the impact of wearing them during an epidemic is not significant. Even at 50% effectiveness in reducing both susceptibility and infectivity and with 50% of the population wearing surgical masks only a 6% reduction in the number of cumulative cases is seen. However, a large proportion of infections are asymptomatic, therefore another mathematical model was studied, by introducing the asymptomatic class to the SEIR model. An improved model (SEIAR)^17^ was studied to analyze the influence of wearing masks on the final size and the basic reproduction number of H1N1. This model shows that, in the absence of facemasks wearing, the cumulative number of infections will be 73%. To demonstrate the effect of the asymptomatic class in the model, numerical simulations of the model were studied presuming that 10%, 25% and 50% of the population wear them under various levels of effectiveness of masks in reducing susceptibility and infectivity. The more effective the masks are and the more they are worn, the more quickly final size will be decreased. In detail, when the proportion of people wearing facemasks is 10%, 25%, and 50%, the final size of H1N1 infections will be reduced by 17%, 28%, and 35%, respectively. Wearing masks can effectively decrease the final size of the infected population. When influenza outbreaks occur, it is almost ineffective to wear facemasks only for infected individuals. In order to effectively reduce infection, all people need to wear facemasks. There is no significant reduction of the final size if only infected individuals wear facemasks. It is important for susceptible, exposed, symptomatic and asymptomatic individuals to wear facemasks and there is a 70% reduction in the final size.

## Discussion

The protective effect of facemasks is determined by the transmission blocking potential of the mask material and the degree of adherence to proper wearing of masks. It also depends on the transmission characteristics of the virus. Most of the studies we reviewed were based on transmission of influenza and influenza like illnesses with few observational studies about the SARS virus. Our study was focused on the usage of surgical/cloth masks in the general population setting.

Experimental studies establish varying degrees of outward protection of masks, indicating that masks worn by infectious subjects (both symptomatic and asymptomatic) will reduce the transmission of infection in the population. The study by van Der Sande et al (2008)^13^, elucidates that masks offer better inward protection than outward protection. Unlike the current guidelines by most countries which assert more on the usage of masks by symptomatic people, the protective effect of masking of healthy individuals in a population has been underscored.

Three of the randomized controlled studies done in community setting suggested significant benefit of face mask usage in reducing the transmission of influenza like illness. None of these studies was however done on a large population. There were no randomized controlled trials done on a larger population for mask usage. Two critical points were also observed in these trials. The benefit of mask usage by the community depends on the time of initiation of usage of masks and the degree of adherence to it. There was much greater advantage when mask usage was started early i.e. right at the beginning of a community infection outbreak, compared with later when secondary transmission might have happened, and more household members may already have been infected by the time of mask adoption. A much greater benefit was also seen when higher percentage of population wore masks. According to the health belief model^18^, adherence with preventive measures increases with higher perception of risk and that is expected to significantly increase during a pandemic. The observational studies done mostly during the SARS epidemic give us a better insight into this behavioral aspect. During the height of the SARS epidemic of April and May 2003 in Hong Kong, adherence to infection control measures was high; and nearly 76% of the population wore a face mask.^12^ Wearing a mask can also raise awareness of the infection risk and the importance of other non-pharmaceutical measures like more frequent hand-washing and social distancing. A face mask may also reduce contact transmission by preventing wearers from touching their mouths or noses with their hands or other objects potentially contaminated with virus. The limitation though is a false sense of security that can lead to poorer compliance with other methods like hand washing.

The transmission characteristics of a virus also plays an important role. Mask usage is practically the only way to prevent aerosol transmission, which may cause the most severe cases of respiratory infections like influenza and Covid-19. Hand washing and social distancing can largely prevent transmission by contact and droplets, but these methods are much less effective against aerosol transmission. The population mask approach is also more beneficial when a disease gets transmitted by asymptomatic carriers. Many SARS-CoV-2 infections are transmitted by people who are asymptomatic.^19^ The length of this asymptomatic period for SARS-CoV-2 infections is estimated to vary from 3-24 days.^20^ This characteristic is unique to SARS-CoV-2 vis a vis other respiratory viruses including SARS and may be reason enough for the entire population to mask up.

In conclusion, Facemask usage by the general population is vital in the prevention of a respiratory virus with unique transmission characteristics as the SARS-CoV-2. While the first priority for facemask usage has to be given to healthcare workers, the general population also needs to adopt masks, as soon as possible. In view of the likely paucity of surgical masks, the community can use homemade cloth masks with adequate precautions of hygiene. The home-made masks still confer a significant degree of protection, albeit less strong than surgical masks or N95/ FFP2 masks. Homemade masks however would not suffer from limited supplies. Even if the transmissibility of the virus reduces by a small fraction due to masking, we will be able to reduce the exponential rate at which the virus has been spreading. It may not mitigate the pandemic but significantly ‘flatten the curve’ and give us more time for the pharmaceutical interventions to take over.

## Data Availability

It is available on request

